# A Mesoscale Agent Based Modeling Framework For Flow-mediated Infection Transmission In Indoor Occupied Spaces

**DOI:** 10.1101/2022.05.20.22275409

**Authors:** Debanjan Mukherjee, Gauri Wadhwa

## Abstract

The ongoing Covid-19 pandemic, and its associated public health and socioeconomic burden, has reaffirmed the necessity for a comprehensive understanding of flow-mediated infection transmission in occupied indoor spaces. This is an inherently multiscale problem, and suitable investigation approaches that can enable evidence-based decision-making for infection control strategies, interventions, and policies; will need to account for flow physics, and occupant behavior. Here, we present a mesoscale infection transmission model for human occupied indoor spaces, by integrating an agent-based human interaction model with a flow physics model for respiratory droplet dynamics and transport. We outline the mathematical and algorithmic details of the modeling framework, and demonstrate its validity using two simple simulation scenarios that verify each of the major sub-models. We then present a detailed case-study of infection transmission in a model indoor space with 60 human occupants; using a systematic set of simulations representing various flow scenarios. Data from the simulations illustrate the utility and efficacy of the devised mesoscale model in resolving flow-mediated infection transmission; and elucidate key trends in infection transmission dynamics amongst the human occupants.

## 1 Introduction

Viral epidemic outbreaks have posed major public health concerns in recent times. The currently ongoing Coronavirus Disease 2019 (or Covid-19) pandemic caused by the SARS-CoV-2 virus (and its variants) is perhaps the most severe of these outbreaks, having led to multiple waves of cases and large-scale fatalities, and a massive worldwide public health and socioeconomic burden^[1]^. Other recent known outbreaks of global significance include the SARS outbreak in the early 2000’s, the H1N1 influenza outbreak in 2009, the MERS-CoV outbreak originating in 2012, and the Ebola virus outbreak in 2014 ^[23,53,13]^. When counted with multiple dispersed influenza outbreaks, a significant number of these diseases involve respiratory system infections with documented airborne transmission modes. Mitigation of severe public health impacts of such outbreaks necessitates a comprehensive understanding of infection transmission and control measures; with a focus on airflow-mediated transmission. This is particularly critical for planning operations in human occupied indoor spaces due to the high potential of infection spread, as reported in a large number of studies^[40,42,58,45,44]^. Transmission of infections like SARS-CoV-2 occurs through respiratory ejecta particles, which can be mediated by transport of such particles by background air flow - leading to *fomite* transmission modes via direct contact; *short-range* transmission modes via large respiratory droplets; and *long-range* transmission modes via aerosolized small/micro-particles^[30,25,71]^. For indoor occupied spaces, this is further influenced by factors such as ventilation, filtration, and occupancy density^[33]^. Together, these aspects motivate the need to advance our understanding of flow-mediated infection transmission in human occupied spaces. With rapidly increasing high-performance computational and digital data-processing capabilities, computational models can provide a viable approach to address this need, by enabling integration of flow physics models with indoor operations data and disease pathogen biology data^[70]^.

The underlying phenomena for flow-mediated infection transmission in such scenarios comprise an inherently multiscale problem^[46,6]^, which is manifested in corresponding approaches for investigating transmission and control. At the very largest scales, compartmental epidemiological models such as SIR (*acronym for Susceptible-Infected-Recovered*) models are employed to study population level transmission dynamics and control intervention strategies^[2,64,8]^. The subsequent coarse scale models commonly investigate infection transmission at larger building scales, using statistical infection transmission models based on well-mixed airflow assumption^[51,52,62]^. No human occupancy or interaction variables are accounted for in detail at such coarse-scale models. Additionally, the well-mixed airflow assumption is not true in most realistic operational scenarios, and often airflow patterns themselves can yield non-intuitive infection transmission patterns^[44,73]^. Conversely, at the very finest scales of phenomenon being modeled, detailed respiratory flow and airflow physics including pathogen-laden respiratory droplet and ejecta formation^[5,17]^ are incorporated. Recently reported methods in this category also include fully resolved fine scale modeling of fluid-particle interactions, thermal effects, and detailed respiratory particle physics^[37,38,36,39]^. However, these models are computationally expensive, requiring multiple cores of computing resources and long compute times for realistic indoor operational scenarios^[39]^. While they are highly effective in enabling a deeper understanding of disease transmission mechanics, they remain intractable for policy and operation decisions pertaining to infection control strategies and interventions. Computational complexity and expense, and need for specialized compute resources, are identified challenges that can prevent adoption and utility of models for infection control and policy decisions^[31,22]^. This motivates the need for mesoscopic modeling approaches of low computational expense, at the intermediate scales where human occupants and their interactions as well as flow-mediated transmission phenomena are accounted for. Several existing studies have documented active agent-based models for systems comprising socially interacting human individuals. Broadly, active agent-based models have a rich history in describing interacting organism behavior. Popular versions include variants of the Vicsek model^[69,68]^ for active organisms, which have been extensively used to study herding and flocking behavior^[24]^, swarm behavior and dynamics^[61]^, collective motion in cellular colonies^[75]^, and other forms of collective dynamics and decision making processes^[55]^. Similar approaches have been extended to human interaction dynamics in form of the Social Force Model^[27]^, the modified Headed Social Force Model^[21]^, and several variations thereof^[74]^, which have been extensively used to study pedestrian dynamics and interacting social behavior^[11]^. Specifically, for infection transmission applications, there have been several attempts using active agent-based models to investigate dynamic transmission and exposure risk: for example during air-travel and airplane boarding and deplaning^[48,49]^; in a crowded supermarket^[26]^; and in pedestrian queues^[18]^; amongst others. In a series of recent works^[37,38,36,39]^ agent based human dynamics models have been coupled with fully resolved fluid-particle interaction models, involving two-way coupling of flow variables and particle transport variables between fluid grid and human agents. These models have been employed to study infection transmission in a variety of scenarios including sneezing in an airplane cabin^[38]^, transmission on a subway station^[38]^, and transmission in a fast-food restaurant^[39]^.

Existing agent-interaction based models used in studying infection transmission either: (a) do not incorporate flow physics and respiratory particle/droplet transport information; or (b) involves fully resolved fine-scale flow-particle and agent interactions, leading to substantial computational expense and complexity. Approaches in category (a) remain insufficient at properly resolving flow-mediated infection transmission - which is critical for respiratory infections such as the Covid-19 pandemic^[30,71]^. Non-uniformity in flow-patterns in indoor airflow is widely acknowledged; yet most agent-based simulations incorporate probabilistic infection risk models which rely heavily on the assumption of well-mixed airflow^[18]^. Consequently, several critical factors such as ventilation, air changes per hour (ACH), airflow isolation strategies, occupancy density and occupancy patterns cannot be comprehensively accounted for in infection transmission and control investigations, thereby potentially leading to sub-optimal infection control strategies and interventions. Conversely, approaches in category (b) above, lead to simulations spanning many millions of degrees of freedom, requiring multiple hours or days of simulation times on many-core machines, for a few minutes of physical time for realistic indoor occupied spaces. These factors further substantiate the need for models at the meso-scale, where model complexity and computational expense remains reasonable for adoption for control interventions, operational decisions, and policy decisions that may involve rapid assessment of multiple *‘what if’* scenarios.

Here, we address this aforementioned modeling need by devising a mesoscopic computational modeling framework that integrates active-agent based human occupant interaction models with a customized semianalytical flow physics based respiratory particle transport model; and, thereby, enables characterization of flow-mediated transmission amongst socially interacting occupants in an indoor space. For the agent-based approach, we use the original formulation of the Social Force Model. Here, we focus on demonstrating the model development in Sections 2 and 3; outlining algorithmic details in Section 4; demonstrating validated implementation in Sections 5.1 and 5.2; and illustrating the utility and efficacy of our model in characterizing flow-mediated infection transmission using a case-study in Section 6. We will not outline details of obtaining full-scale CFD models of building airflow as function of ventilation to keep with the scope of this study; and refer to our prior works^[44,73]^ for further details on this separate aspect. All models have been implemented in custom computer codes developed in Python using the scientific computing stack (NumPy, SciPy); and agent-data post-processing have been conducted using custom scripts based on the open-source Visualization Toolkit (VTK) library.

## 2 Mathematical description of social force models

### 2.1 Basic premise of the model

We will state the mathematics of the social force models, following the formulations and details in^[27]^, for a system of *N* active human pedestrians (each indexed by *i* ∈ [1, *N*]). We assume that each pedestrian agent *i* has a desired velocity ***<u>w</u>***_*i*_ with which the agent intends to travel within the domain of movement. The fundamental premise of agent-based social interaction models is that changes in the desired velocity ***<u>w</u>***_*i*_ are influenced by a combined set of phenomenological forces:

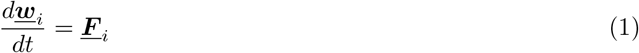

Specifically for the classical formulation of social force model^[27]^, this includes a collection of forces that determine collision avoidance, social affinity, and navigation to a specific target. To describe these further, we introduce a second agent velocity - the actual velocity ***<u>v</u>***_*i*_ - which is the true velocity with which the agent physically moves, and therefore changes its location ***<u>x</u>***_*i*_ as:

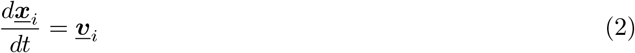

Assuming further that each agent can only achieve a maximum achievable speed *v*_*max*_, we define a function *g*(*x*) as follows:

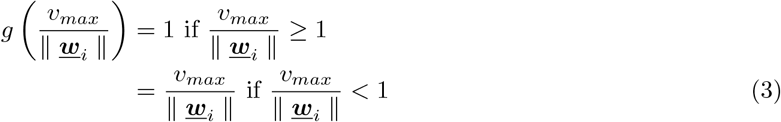

We then relate the actual and the desired velocities as follows:

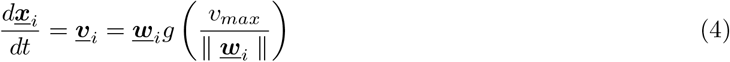

Specifically, if the actual maximum possible speed of motion is less than the desired velocity, then the agent can achieve their desired velocity; otherwise the actual velocity is a scaled version of their desired velocity. This accounts for constraints on motion based on realistic human locomotion biomechanics considerations. For the social force model as proposed in^[27]^, the combined social force ***<u>F</u>***_*i*_ and the resulting change in agent position is stated as follows:

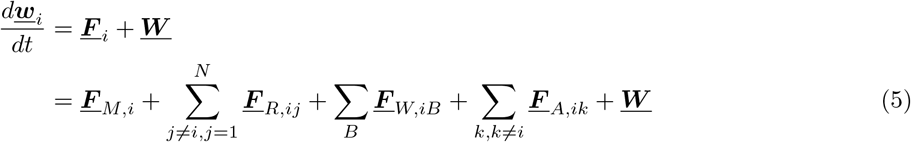

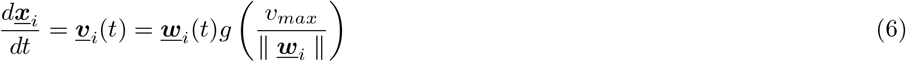

In Equation 5, ***<u>F</u>***_*M,i*_ denotes a directional force to maneuver in a desired direction; ***<u>F</u>***_*R,ij*_ denotes a collision avoidance force between agents *i, j*; ***<u>F</u>***_*iB*_ denotes a wall collision avoidance force between agent *i* and wall *B*; and ***<u>F</u>***_*A,ik*_ denotes a social affinity force between agent *i* and agent or wall or spatial location denoted by *k*. Formulation of each force contribution is outlined in the subsequent sections. The term ***<u>W</u>*** is a stochastic term modeling fluctuations in agent dynamics. The stochastic fluctuation term ***<u>W</u>*** is commonly modeled as a Gaussian process with a zero mean, similar in form to the fluctuations in Langevin dynamics or active-agent Vicsek type models^[59,68]^.

### 2.2 Directional forces for each agent

Each agent is assumed to navigate towards a sequence of destinations ***<u>d</u>***_*α*_ with *α* ∈ [1, *K*] and *K* being the total number of target destinations that the agent will be likely navigating. Using this, we define a desired navigation vector 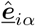 for an agent from its current location ***<u>x</u>***_*i*_ to the *α*’th destination point as follows:

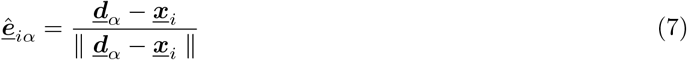

Assuming that the agent desires to get to ***<u>d</u>***_*α*_ with velocity 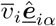, but is only able to navigate as per the actual velocity ***<u>v</u>***_*i*_, we can state a direction force as follows:

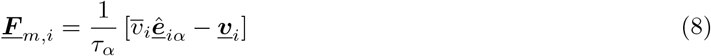

with *τ*_*α*_ being a relaxation time to account for pedestrian navigation velocity changes due to any acceleration or deceleration originating from social interactions. Here, *τ*_*α*_ physically plays the same role as particle momentum response time in particle-laden flows and discrete element methods^[15,47]^; meaning that a low *τ*_*α*_ will enable agent to quickly approach the desired velocity; while higher *τ*_*α*_ will make agent motion more sluggish.

### 2.3 Pedestrian collision avoidance

When distance between two pedestrian agents becomes closer, territorial behavior of agents will lead to momentum changes to ensure collision avoidance. This is accounted for using a potential based repulsion force ***<u>r</u>***_*ij*_ based on a repulsion potential function ℛ defined as follows:

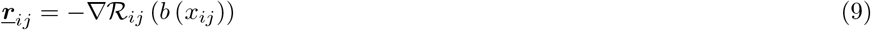

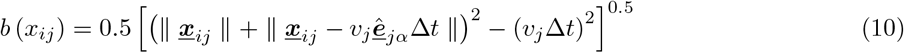

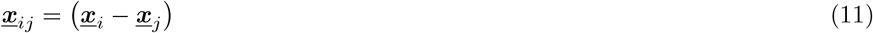

In reality, this force needs to be scaled based on agent direction 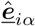, as the change in momentum will differ when a neighboring agent *j* is located right in front of agent *i* compared to when located behind agent *i*. This scaled force is stated as follows:

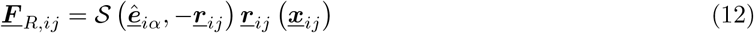

where the scaling function 𝒮 is defined for any given pair of vectors ***<u>e</u>*** and ***<u>f</u>*** as follows:

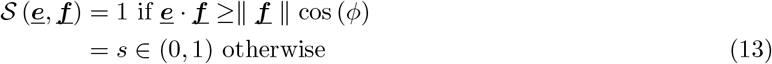

and the angle 2*ϕ* represents the pedestrian agent perception angle of sight.

### 2.4 Wall collision avoidance

Realistic scenarios of agent dynamics must account for collision avoidance with walls and boundaries of spaces, as well as other indoor structures that substantially modify the human occupied space (for example, large furniture, space barriers etc.) We follow a potential based repulsion force formulation to account for momentum changes due to this collision avoidance as well. For any boundary wall or structure wall ℬ we identify ***<u>p</u>***_*Bi*_ - the point on ℬ that is the closest to pedestrian *i*. Using this point coordinate, we can now state the wall collision avoidance force ***<u>F</u>***_*W,iB*_ based on a wall collision potential 𝒲 as follows:

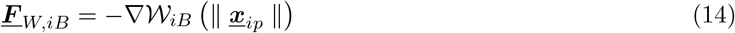

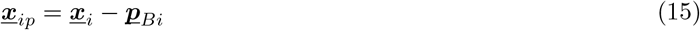

We note that the potential force formulations in Equations 12 and 14 have similarities to potential-based forces in molecular dynamics, Langevin dynamics, and discrete element mechanics models. However, for maintaining consistent collision avoidance, and ensuring no spurious acceleration is allowed, the potentials must have a monotonic behavior with approach distance for both cases.

### 2.5 Social affinity interactions

Human agent momentum changes may occur due to attraction or affinity towards other agents, leading to the formation of socially friendly groups of individuals, or affinity towards other non-human targets in the space (for example, displays in a museum etc.). To account for this, we employ a similar potential based approach as in Equations 12 and 14. Specifically, we state the attraction force ***<u>a</u>*** based on an affinity potential A between an agent *i* and another agent or object *k* as follows:

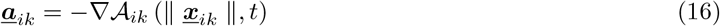

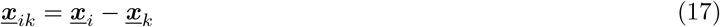

and we scale this force based on location of the agent or object *k* with respect to the agent *i* (perception or line of sight) using the same scaling function S as described in Equation 12 as follows:

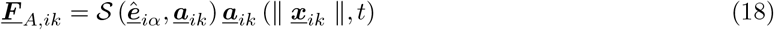

where, 𝒮 retains the same definition as in Section 2.3. The affinity potential 𝒜 is allowed to vary over time for this model, to enable changes in varying affinity towards a specific individual agent or object of interest. Similar to ℛ, and 𝒲, monotonic behavior with approach distance for the functional form of is 𝒜 desirable to avoid spurious oscillations in agent dynamics.

## 3 Flow-mediated infection transmission model

### 3.1 Semi-analytical model for droplet dynamics in airflow

In this computational framework, we account for the airflow-mediated multiphysics phenomena involved in respiratory droplet dynamics from infected host to susceptible individuals through a semi-analytical approach based on a set of coupled ordinary differential equations describing the dynamics of an individual evaporating droplet. The general approach is outlined conceptually in Figure 1. We consider the dynamics of a respiratory droplet of diameter *D*_*p*_ = 2*R*_*p*_ moving in indoor flow with background air density *ρ*_*f*_ and viscosity *µ*, and velocity ***<u>u</u>*** (interpolated at the individual droplet location). Assuming individual droplet size is small in comparison to larger indoor flow length-scales (that is, small particle slip-velocity Reynolds numbers, as well as small Biot numbers for temperature variations), to a first-order approximation we can write the coupled set of ODEs for individual droplet as follows:

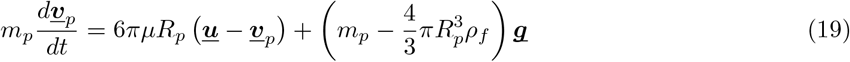

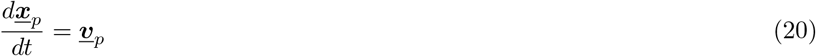

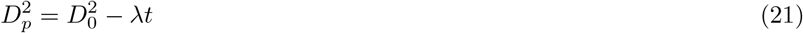

where Equation 19 accounts for drag from the surrounding flow, gravity and buoyancy effects; and Equation 21 describes droplet diameter change based on evaporative mass transfer theory as outlined in several previous works^[15,43]^. While more detailed droplet dynamics and evaporation models can be formulated as outlined in several other studies^[57,4,10]^, here the simplified equations above provide a robust approach to semi-analytically integrate flow-mediated droplet multiphysics into social interaction driven agent-based modeling. The parameter *λ* is derived from evaporative mass transfer considerations, and effectively determines the droplet desiccation lifetime to a certain final droplet diameter *D*_*f*_ stated as follows:

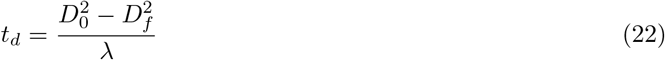

**Figure 1:**
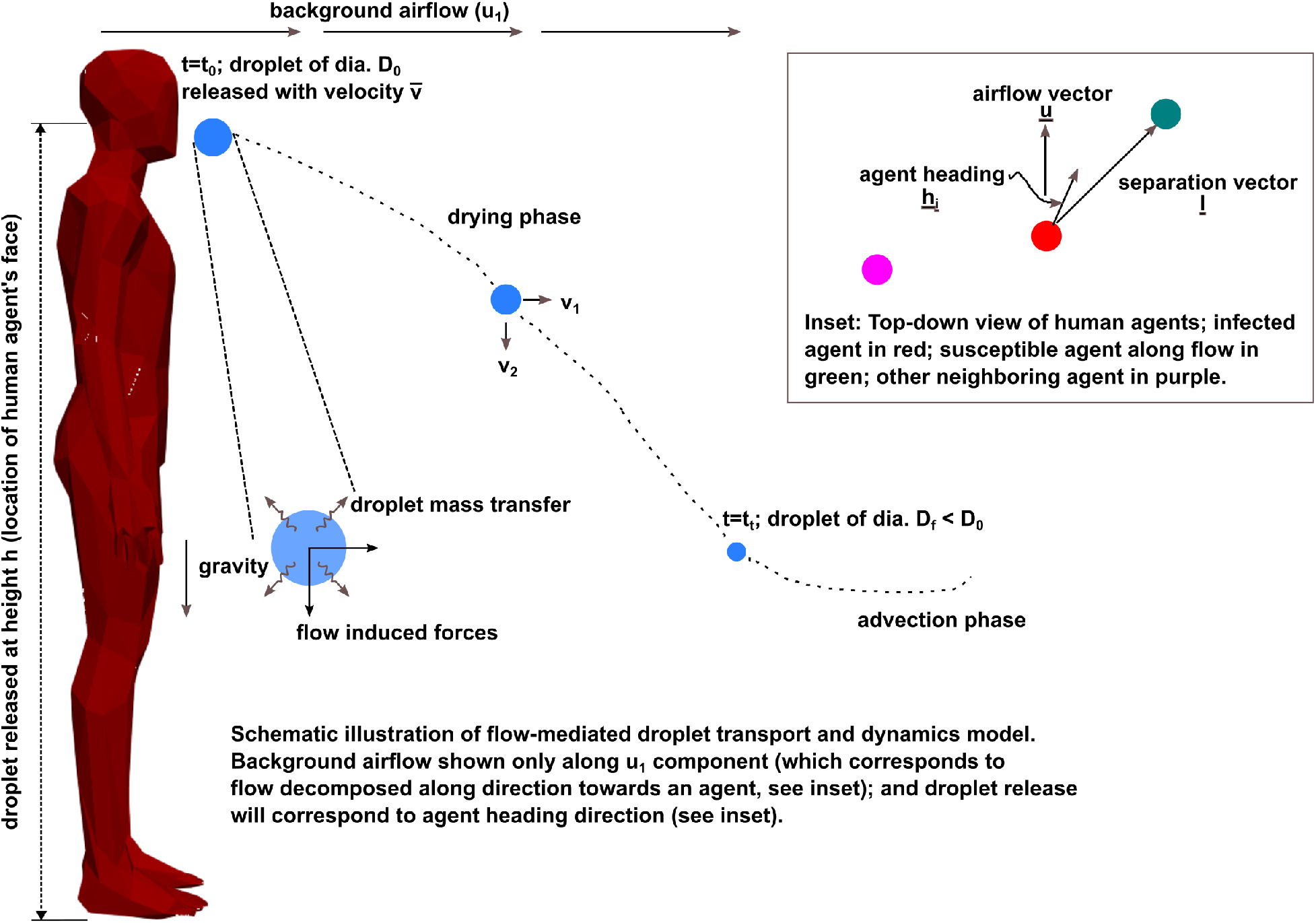
A visual illustration of the key aspects of the proposed agent-based flow-mediated infection transmission model; identifing the flow physics aspects for droplet transport, and (in inset) the scheme used for agent-based exposure identification based on droplet dynamics.

It can be shown that *λ* can in turn be more precisely stated in terms of specific droplet mass transfer quantities^[15]^ as follows:

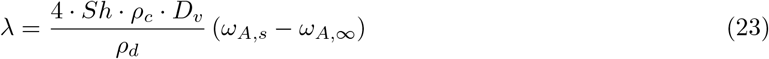

where, *D*_*v*_ is the mass diffusion coefficient of droplet vapor; *ω*_*A,s*_ is the mass fraction of vapor corresponding to the droplet liquid evaluated at the droplet surface; *ω*_*A,∞*_ is the mass fraction of the same vapor at freestream; *ρ*_*c*_ is gas density at free-stream; and *ρ*_*d*_ is the droplet liquid density; and *Sh* is the droplet mass transfer Sherwood number, commonly expressed in terms of the Ranz-Marshall correlation stated as follows:

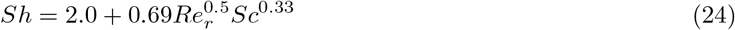

for *Re*_*r*_ being the droplet slip-velocity Reynolds number, and *Sc* being the droplet fluid Schmidt number. Due to the continuous reduction in droplet diameter, we must ensure that *D*_*p*_ > 0 physically for all scenarios; and that at the limit of *D*_*p*_ ≈ 0 we end up at the tracer-limit for the droplet, such that droplet velocity will be the same as background flow velocity. In reality for respiratory droplets, the diameter will reduce to a minimum *D*_*f*_ > 0 where droplet size does not reduce further, and instead a nucleus of proteins, salts, and other biochemical by products in respiratory ejecta is formed^[50,71]^. This is illustrated in Figure 1 where individual droplet undergoes a continuous *drying phase* until a transition time *t*_*t*_ where *D* = *D*_*f*_ and thereafter the droplet is in an *advection phase*. Restating Equations 19 and Equations 21 in terms of the momentum response time *τ*_*p*_, we obtain the following:

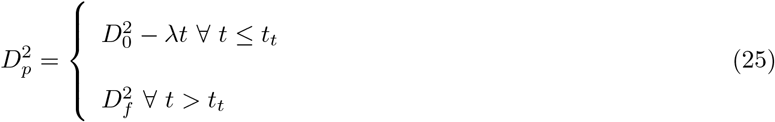

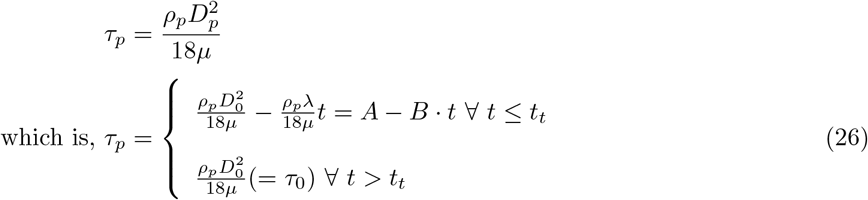

such that we get:

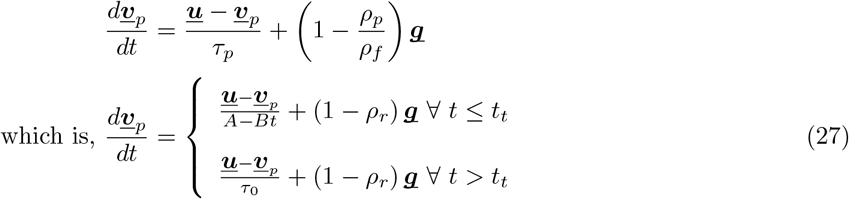

Using the coordinate system notation as outlined in Figure 1, we can now analytically integrate the stated ODEs for droplet velocity and trajectory, for the scenario where a human agent with their face located at height *h* from the ground releases a respiratory droplet at a velocity 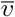 in the direction of its own heading. At the transition time when the droplet reaches the nucleus diameter of *D*_*f*_, we assume that the droplet coordinates are given by 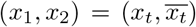 and velocity 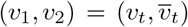. The resulting expressions for velocity components *v*_1_, *v*_2_ and trajectory coordinates *x*_1_, *x*_2_ in the height-wise cross-sectional plane of the human agent, for the *drying phase* during *t* ≤ *t*_*t*_ can be stated as follows (*detailed derivation excluded*):

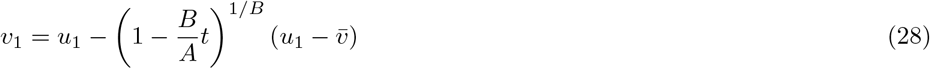

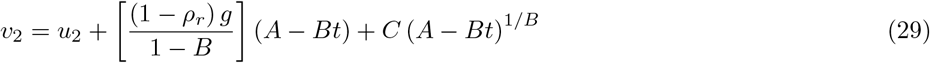

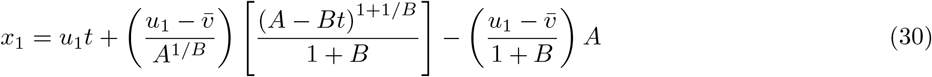

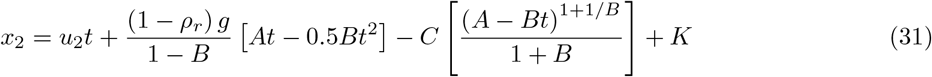

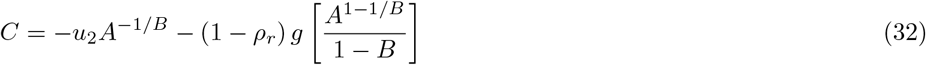

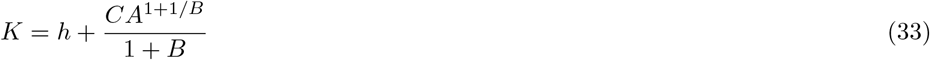

Following the same analytical derivation, but for the non-drying *advection phase* at *t* > *t*_*t*_ and with *D*_*p*_ = *D*_*f*_, we obtain the expressions for *v*_1_, *v*_2_ and *x*_1_, *x*_2_ as follows:

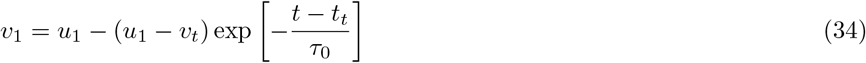

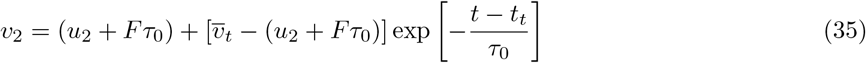

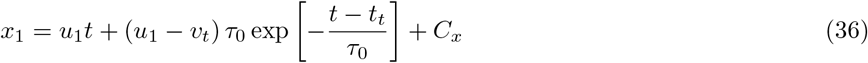

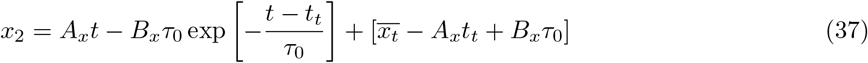

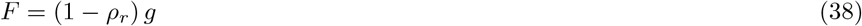

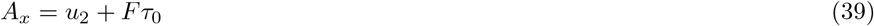

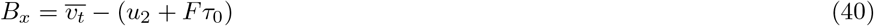

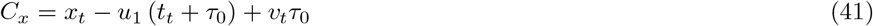

The Equations 28-37 together constitute a procedure to obtain the droplet dynamics variables ***<u>v</u>***_*p*_, ***<u>x</u>***_*p*_, *D*_*p*_ as a function of air-flow velocity ***<u>u</u>***, other flow-related variables, and human agent variables (such as *h* and 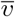). For the purpose of the rest of the computational framework description, we will identify this aggregate of integrated equations as an overall respiratory droplet physics model `RD` such that: 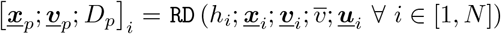. We use this notation to devise an infection exposure and transmission model next.

### 3.2 Infection exposure and transmission potential

The fundamental premise of infection transmission in the scenarios discussed here is that a non-infected but susceptible individual can be infected by an infected individual when virus-laden respiratory carrier particles from the latter are exposed to the former. This typically constitutes two components. First, an effective contact between infected and susceptible agents is evaluated, and an equivalent exposure to infection is identified. Second, the contact and exposure can be used to further define an infection probability. Contact and exposure will further depend upon pathogen type. Certain infections (like Ebola^[14]^) require actual physical contact for transmission; some pathogens require short-range contacts for transmission; and some remain viable and suspended in air for sufficiently long durations to enable both short-range and long-range transmission (such as Covid-19 ^[30]^). Existing studies typically define contact based on a simple cutoff distance thresholding between an infected agent and a susceptible agent. Here, we use the respiratory droplet physics to inform a flow-mediated exposure model. The schematic of this approach is outlined in Figure 1. We consider an infected agent (*marked in red*), and two agents around them, with one susceptible agent (*marked in green*). We define a local airflow ***<u>u</u>***, and infected agent heading direction ***<u>h</u>***_*i*_, and project ***<u>u</u>*** along the vector joining the infected and susceptible agent locations ***<u>l</u>*** to obtain *u*_1_ and *u*_3_ (orthogonal to *u*_1_ such that *u*_2_ is out of the plane of the agent). The respiratory droplet release velocity is then similarly decomposed as 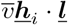 relative to the agent (that is, infected agent velocity is added to the droplet exit velocity). These inputs are fed into 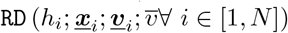 to obtain droplet location ***<u>x</u>***_*p*_ and distance from infected agent ‖ ***<u>x</u>***_*p*_ −***<u>x</u>***_*i*_ ‖; when the model `RD` is evaluated for a threshold integration time *T*_*c*_. The threshold time *T*_*c*_ can be set based on inhalation-exhalation timescales of normal human breathing; and can be typically in the range of 2-4 seconds. If a susceptible agent at the instance of exposure evaluation is within this droplet traversal distance, we identify an exposure event, and update contact and exposure data for the susceptible agent. When multiple infected and susceptible agents are present, this computation can be conducted pairwise for each pair of susceptible and infected agents.

## 4 Algorithmic aspects for simulation framework

### 4.1 Structural boundaries

The agent dynamics simulations occur across computational domains that are 2-dimensional top-down views of the occupied spaces they are navigating. The boundaries of these spaces are composed using polyline segments, denoted in 2-dimensional space using the end coordinates of the segments: ℬ = {***<u>x</u>***_*b*1_, ***<u>x</u>***_*b*2_}, for each wall or boundary object ℬ. The same is used for identifying any indoor barrier, furnishing, or structural boundary. Target destination points ***<u>d</u>***_*α*_ can then be set as points along the line segment connecting ***<u>x</u>***_*b*1_ and ***<u>x</u>***_*b*2_: for example ***<u>d</u>***_*α*_ = ***<u>x</u>***_*b*1_ + *d* · (***<u>x</u>***_*b*2_ − ***<u>x</u>***_*b*1_) / ‖ ***<u>x</u>***_*b*2_ − ***<u>x</u>***_*b*1_ ‖ with *d* being a location parameter. Additionally, closest point ***<u>p</u>***_*Bi*_ for determining wall collision avoidance for agent *i* can be determined based on standard geometric calculations of distance of a point from a line segment^[20]^. For infection exposure estimations, it is important to identify whether an infected and a susceptible agent are on the same side or opposite sides of a boundary segment. This is identified, by checking whether the line segment joining the agents intersects a boundary segment; and by marking agents on same or opposite side based on whether intersection has occurred. We note that, it is possible for flow and transmission to occur across a boundary segment, especially if the segment is not an impervious structural barrier, if the segment has a window or ventilation passage, or if overall building ventilation can move flow across the segment. Lastly, the actual boundary wall segments can be either manually specified, or obtained directly from architectural drawings, floor plans, and room or building layout data, using techniques similar to those demonstrated in our prior work^[44]^.

### 4.2 Occupancy pattern initialization

We use a discrete representation of indoor space occupancy based on our previous work^[73]^, where occupancy is outlined in terms of vector 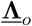 comprising the location ***<u>x</u>***_*k*_ of each individual agent or occupant:

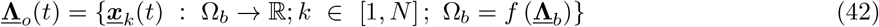

with *N* being the number of agents; Ω_*b*_ denoting the interior domain of the occupied space; and 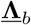 denotes the parameter vector containing data on the domain geometry including built space layout, floor-plan, and dimensions. Once initialized with an occupancy pattern 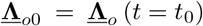, the social force model will yield subsequent outputs of 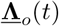. Here, using the parameters in 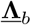, we use a random sequential addition process^[72,66]^ to generate an initial 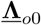 configuration of agents represented geometrically as a circular disk with a specified radius of influence. The geometry parameters 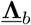 define rectangular rooms, corridor spaces etc. We note that random sequential addition is suitable and robust for initializing low to moderate occupancy densities (*N/*vol (Ω_*b*_)), which was the focus here for model development. For more dense and crowded occupancy scenarios, geometric tessellation based approaches^[16]^ or dynamic algorithms such as Lubachevsky-Stillinger algorithms^[41,65]^ will be necessary.

### 4.3 Target and destination scheduling

Specification of a sequence of targets and destinations ***<u>d</u>***_*α*_ (as stated in Section 2.2) is a critical piece of the algorithmic outline; wherein a planned sequence of point coordinate inputs are needed. This sequence: (a) can be manually assigned for simple simulation scenarios; (b) can be obtained using detailed path planning simulations using techniques similar to Dijkstra’s algorithm^[19,26]^; or (c) can be obtained from operational data for the indoor space. Here, to keep with the scope of method development, we employ a more direct approach based on agent location. Specifically, doors in the space layout are input as line segments similar to boundary segments ℬ in Section 4.1 above; and at each time closest point on a door segment from a given agent is computed, and updated into the series ***<u>d</u>***_*α*_ for *α* ∈ [1, *K*]. We note that, these line segments can be other locations in addition to doors as well; however, for our case study presented in Section 6 we use these segments only for doors since the scenario models all occupants exiting a building.

### 4.4 Numerical implementation

Here, we devise a generalized numerical implementation using the social force model position and velocity updates based on an explicit Runge-Kutta (RK) method with *s* stages; and the overall algorithmic framework has been illustrated in Algorithm 1. Explicit RK methods are chosen here for simplicity of implementation, and ability to achieve customizable high-order accuracy. For model implementation for this study, we have chosen the popular 4-stage Runge-Kutta integrator (RK4). Specifically, since the social force dynamics does not depend upon background airflow information, the agent positions and velocities are updated first (using the Runge-Kutta integrator); followed by the interpolation of airflow velocities at agent locations; and subsequently, update of respiratory droplet model to evaluate contact and exposure of a susceptible agent to an infected agent. The choice of potentials ℛ, 𝒲, 𝒜 and requires some numerical considerations, such that the forces remain monotonic in behavior to avoid artificial acceleration and deceleration during agent movement (See also Section 2). As per recommendations in prior studies^[27,21]^, these potentials were chosen as exponential functions. In addition, force computations for collision avoidance and affinity interactions, as well as exposure and contact for infection, will require pairwise checks between agents. For small number of agents (as is the case here) naive pairwise checks for distance between agents can be computed directly. However as the number of agents increases, classic nearest-neighbor based techniques such as cell-lists^[59,47]^ will need to be employed. Our proposed framework adds the contact and exposure time in a pairwise manner for each susceptible agent with respect to each infected agent (see, for example, lines 15-23 in Algorithm 1). We note that for cases where airflow velocity ***<u>u</u>*** is obtained from building scale CFD, or grid-based measurement data, the interpolation procedure in line 17 of Algorithm 1 will involve a grid-based interpolation operation such as binning operations or trilinear interpolations in structured Cartesian grids and *cell walking* algorithms based on triangle barycentric coordinates^[29]^ in unstructured triangular grids. Finally, the presence of the random fluctuation term ***<u>W</u>*** needs to be accounted for in the integration scheme in a suitable manner. Stochastic multi-stage integration schemes based on Ito calculus have been proposed^[63]^ in existing studies, and the algorithm described here (Algorithm 1) can be extended to implement such methods. Alternatively, a simplified approximation can involve scaling the fluctuation contribution at each stage of Runge-Kutta integration to ensure overall fluctuating displacement scales correctly with Δ*t* time-step size, such that mean square displacement is correctly predicted due to fluctuations.

## 5 Verification of model implementation

### 5.1 Verification of social force behavior

We first establish a verification of the social force model implementation with the algorithmic details presented here. For this, we simulate the self-organization behavior of pedestrians traversing along a walkway as detailed in Helbing’s study^[27]^. Specifically, pedestrians enter the walkway at random positions at both ends of a rectangular walkway, and move with desired velocities 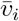 being a Gaussian random variable with mean 1.34 m/s and variance 0.068 m^2^s^-2^. Agent momentum relaxation time *τ*_*α*_ was set to 0.5 s. Collision avoidance potential ℛ was modeled as ℛ(*x*) = ℛ_0_. exp (−*x/σ*), with ℛ_0_ = 2.1 m^2^s^-2^, and *σ* = 0.3 m. Wall collision avoidance potential was modeled as 𝒲(*x*) = 𝒲_0_. exp (−*x/R*), with 𝒲_0_ = 10 m^2^s^-2^ and *R* = 0.2 m. The social affinity potential was set to 0. Agent perception angle of sight *ϕ* was set to 100 degrees; with the scalar scaling constant 𝒮 for the function in the force equations being set to 0.5. Above a certain pedestrian occupancy density, spontaneous formation of lanes are observed as pedestrians try to maintain a uniform walking direction. Helbing’s^[27]^ original study showed that the average number of lanes formed (*N*_*L*_) will scale with the walkway width *W* as *N*_*L*_ ≈ 0.36*W* + 0.59. Results from our implementation are computed and compared against those in ^[27]^ in Figure 2. It is observed that the predicted number of lanes formed obeys the same scaling relation with *W* as reported in the original case-study. Additional visualization of the case of lane formation is presented in form of animations of pedestrian agents provided in the Supplementary Material.

**Figure 2:**
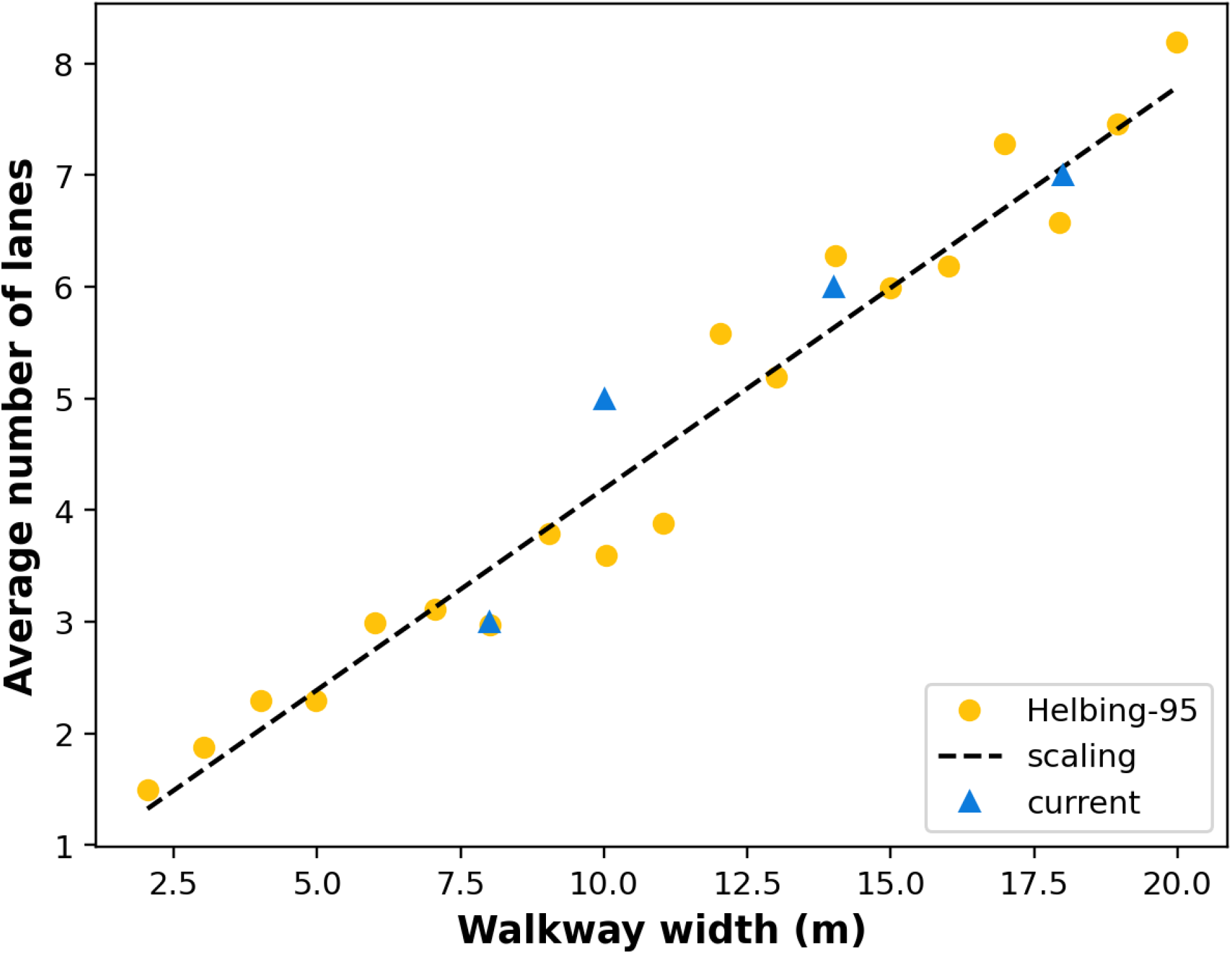
Validation of social force implementation using the lane formation examples stated in Helbing’s original social force model study^[27]^. Helbing’s data, and the proposed scaling relation are illustrated in yellow dots, and solid dashed lines respectively. Current model predictions, represented in blue triangles, line up in accordance with the prior data, thereby establishing validation of current model implementation. See also lane formation animations presented in Supplementary Material.

### 5.2 Verification of infection exposure model

The implementation of the proposed infection exposure and transmission model is validated using a simplified model scenario, involving a group of 100 individual susceptible agents arranged in accordance with a regular Cartesian grid (10 × 10), who are facing an individual agent who is infected (see illustration in Figure 3). A constant airflow field is imposed, and exposure is evaluated based on the modeling approach outlined in Section 4, with any individual who is exposed being marked using a binary indicator to distinguish them from remaining susceptible population (marked in black and white as shown in all illustrations in Figure 3). Several different parameter variations were tested, to verify whether the model captures the exposure behavior in a realistic manner. First, velocity direction was kept fixed in a direction directly facing the susceptible group from the infected individual; and velocity magnitudes were varied from 0.1 m/s to 1.0 m/s. The resulting exposure patterns for increasing velocity magnitudes are presented in Figure 3, panel a. The observations indicate an expected trend where with increasing velocity more agents are exposed, and the exposure spreads to a greater distance away from the infected agent. Second, velocity direction and magnitude were both varied to determine whether exposure trends varied consistently, and resulting trends are illustrated in Figure 3; panel b. Third, the droplet integration threshold time *T*_*c*_ was varied between the typical range of 1 −4 seconds (see Section 3.2), and resulting trends are illustrated in Figure 3, panel c. The illustrations indicate consistent behavior of the exposure trends with both parameter variations; with the integration time *T*_*c*_ showing a significant effect in determining overall exposure patterns.

**Figure 3:**
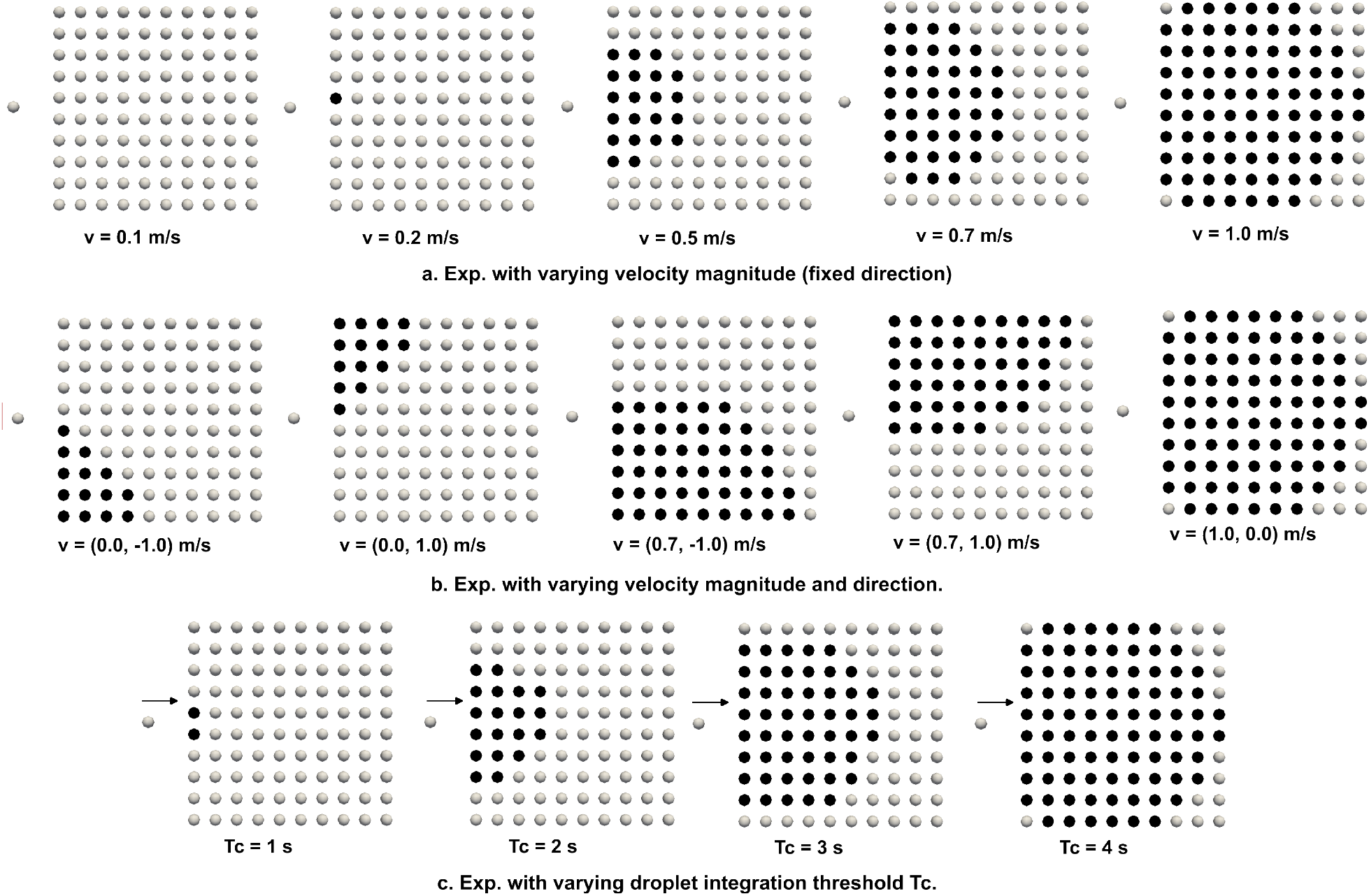
An illustration supporting verification of our proposed infection exposure and transmission model. Exposed individuals are marked in black, while susceptible individuals are marked in white. Panel a. depicts results for exposure with varying airflow velocity magnitude, but fixed direction Panel b. depicts the same for varying velocity magnitude and direction. Panel c. depicts trends in exposure with increasing droplet integration threshold time T_c_.

## 6 Simulation case study for a model indoor space

Having validated the social force and the infection exposure model implementation, we present a simulation case-study to demonstrate our complete computational framework (as outlined in Algorithm 1). For this case-study we consider a representative model indoor space as illustrated in Figure 4, left panel with four rooms connected by corridors towards exits at ends of hallways on both ends. Room dimensions are specified (in meters) in the floor plan layout, and locations of doors are identified in green rectangles. We simulate a specific social dynamics scenario where the building has *N* = 60 occupants (15 occupants in each room) who are trying to exit through the corridor out of the building through one of the exit hallways. This model representation would, for example, correspond to a scenario where individuals are exiting an office building at the end of a business day (amongst other possible scenarios). We manually impose an airflow pattern as demonstrated in Figure 4, right panel. We note that for more realistic simulation models, this airflow pattern can be obtained based on building scale airflow simulation computations or measured airflow data (see also Section 7); which we have not considered here for conciseness of scope of the study. Instead, the imposed simplified airflow pattern enables accounting for sufficient spatial heterogeneity to demonstrate utility and efficacy of our computational approach.

**Figure 4:**
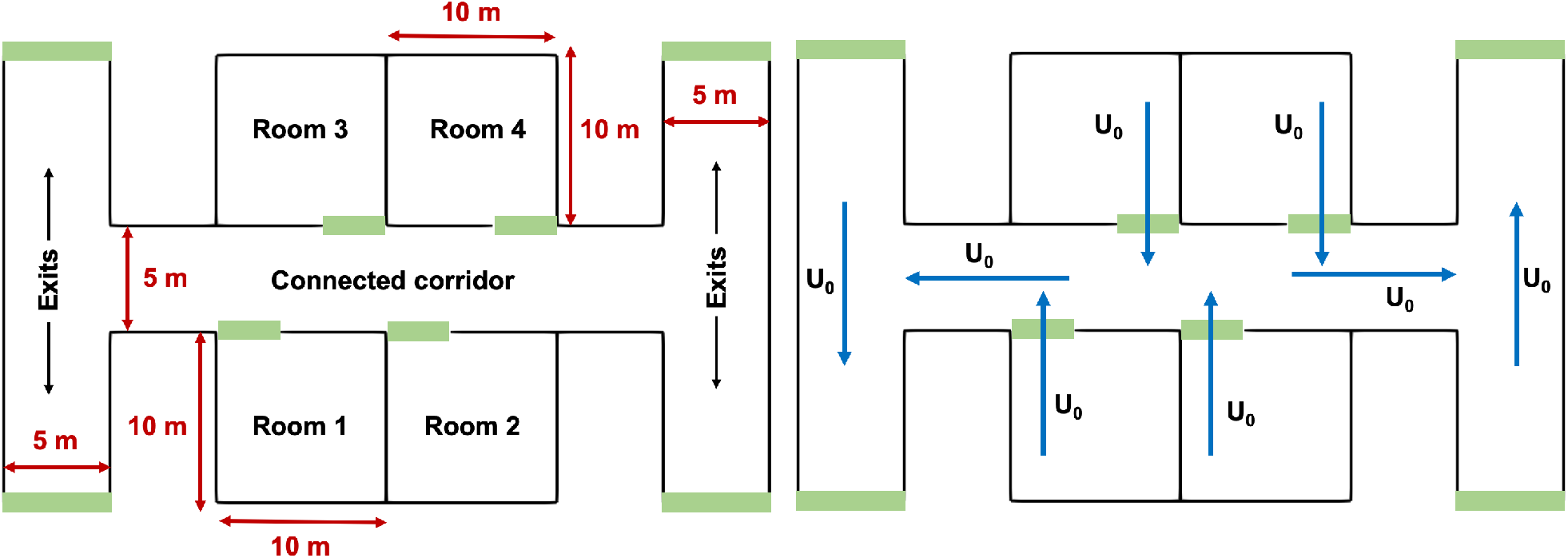
Illustration of the simulation set-up for the case-study outlined in Section 5.2. The indoor space layout and floorplan is shown on the left, with dimensions of each room stated. Simulation set-up comprised 15 agents in each room 1-4, with a total N = 60 agents. Manually specified airflow pattern through the space is depicted on the right, with U_0_ representing a fixed velocity magnitude throughout, that is varied in the simulation study as outlined in Section 5.2.

For the purpose of this study, a series of four different airflow velocity magnitudes (*U*_0_ as per Figure 4, right panel) are considered: 0.1 m/s; 0.2 m/s; 0.5 m/s; and 1.0 m/s. Two different scenarios are explored. In the first scenario, the walls are treated as impermeable structural barriers that partition the indoor space, and airflow does not transmit across the walls. In the second scenario, the walls are treated as structural partitions that guide and direct human occupancy, but are pervious to airflow across (that is, either through windows or open passage of air, or through air transfer across ventilation pathways). To demonstrate that our model captures the influence of flow-mediated transmission as compared to transmission without considering airflow; two additional simulation scenarios are considered. In these, the droplet flow-physics is not integrated, and contact and exposure is defined simply based on a cutoff separation distance threshold of 1.5 m; and 2.0 m as informed by many prior studies on Covid-19 transmission (especially, the commonly used 6 feet or 1.8 m social distancing recommendations)^[30,32]^. The combination of these led to 12 different simulation cases; each of which were numerically integrated using a time-step of 0.25 seconds until the time that all agents effectively reached the exit doorways. For each case, the potentials ℛ and 𝒲 were chosen as described in Section 5.1, while affinity potential A was set to 0. The initial droplet diameter was set to *D*_0_ to 5.0 *µ*m, and *D*_*f*_ set to 2.2 *µ*m, to mimic normal respiratory ejected particles (no coughing and sneezing); the drying coefficient *λ* was set to 0.876 *µ*m^2^/sec; the droplet integration threshold *T*_*c*_ set to 4.0 seconds; and the initial droplet release velocity 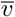 at 40.0 m/s. The resulting agent dynamics, and exposure time for infection were analyzed to demonstrate utility and efficacy of our modeling framework.

Figure 5 depicts a successive series of 5 snapshots (panels a-e) representing the collective dynamics of the *N* = 60 human agents who are moving from their respective rooms towards the exit doorways. Panel f. in Figure 5 represents the effective emergent flow pathlines for the agent collective dynamics, which illustrates the spontaneous formation of an ordered exit flow pattern (indicated by a clustering of pathlines near doors of each room). We note that the number density of agents considered here is much lower than other evacuation scenario simulations reported in literature, and hence the typically observed jamming transition of active agents is not observed here^[68,54]^. In Figure 6, we map out the exposure time for all agents when considering one single infected agent located initially in room 1 (as marked in the figure), along each pathline trajectory. Three scenarios are shown here: panel a. depicts the case for airflow 0.5 m/s with structural walls fully impervious to any flow or transmission across them; panel b. depicts the case for airflow 0.5 m/s with airflow passage and transmission across the wall boundary lines; and panel c. depicts the case for no airflow, but a constant threshold radius of 2.0 m (a little over the 6 feet/1.8 meters physical distance recommendation). The effect of airflow is observed in terms of a greater spatial extent of exposure in panel b., followed by panel a., and panel c. respectively; while panel c. shows the greatest magnitude of exposure owing to continuous contacts within the fixed radius threshold irrespective of airflow. Consideration of separation distance alone without airflow, leads to a spatial localization of estimated exposure to infection, thereby poorly resolving long-range infection transmission, as well as short-range transmission depending upon the complexity of airflow patterns. This is further illustrated in Figure 7, where we present a statistical distribution (including the mean values) of exposure for all agents for all the 12 simulation scenarios considered. Panel a. presents the exposure data samples for all agents from scenarios with no flow or transmission across structural walls; and panel b. presents the exposure data for all agents for scenarios where flow/transmission across the boundary lines was allowed. In both cases, mean exposure generally increased with increasing *U*_0_ magnitudes; with panel a. depicting a narrow banded sample distribution and lower mean, owing to the fact that only local flow effects are considered (no transmission across walls). Conversely, panel b. depicts a wider banded sample distribution, and higher mean compared to panel a. A greater extent of airflow transports infectious particles to more susceptible agents within the critical threshold time *T*_*c*_ = 4.0 seconds, leading to higher extent of long-range transmission; while the constant cutoff case simply captures the localized short-range effects. The simulation data also provide some insights into the dynamics of transmission and exposure; as illustrated in the variation of exposure over time for all *N* = 60 agents (and the mean exposure across all agents) in Figure 8. The left column presents all cases with changing flow magnitude *U*_0_ for no transmission across walls; while the right column presents the cases with transmission across walls (with *U*_0_ magnitudes increasing from top to bottom panel). In all cases, exposure trends show an initial rapidly rising phase, followed by a slow varying phase reaching nearly a plateau. As expected, the obtained exposure dynamics trends is significantly mediated by airflow; as well as the nature of human agent dynamics. Additional visualization of agent dynamics, and exposure behavior is presented in form of a series of animations provided in the Supplementary Material.

**Figure 5:**
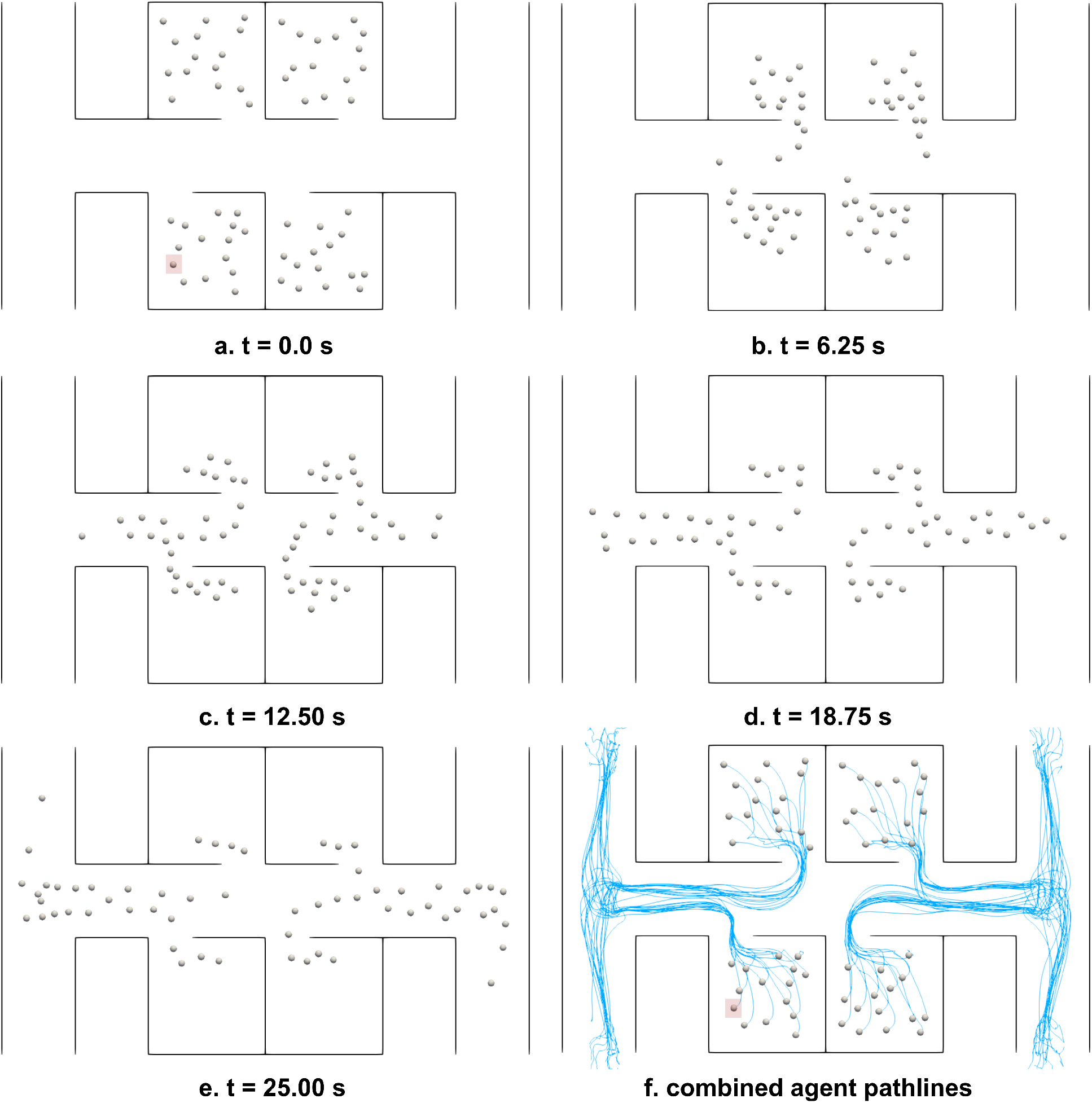
Illustration of interacting human agent dynamics through the occupied space for the simulation case-study. Panels a.-e. depict the agent positions at successive instances in time, as they agents navigate from their respective rooms to the exit doorways. Panel f. presents the emergent flow pathlines from the collective agent dynamics. The infected agent is marked in red square on panels a. and f.

**Figure 6:**
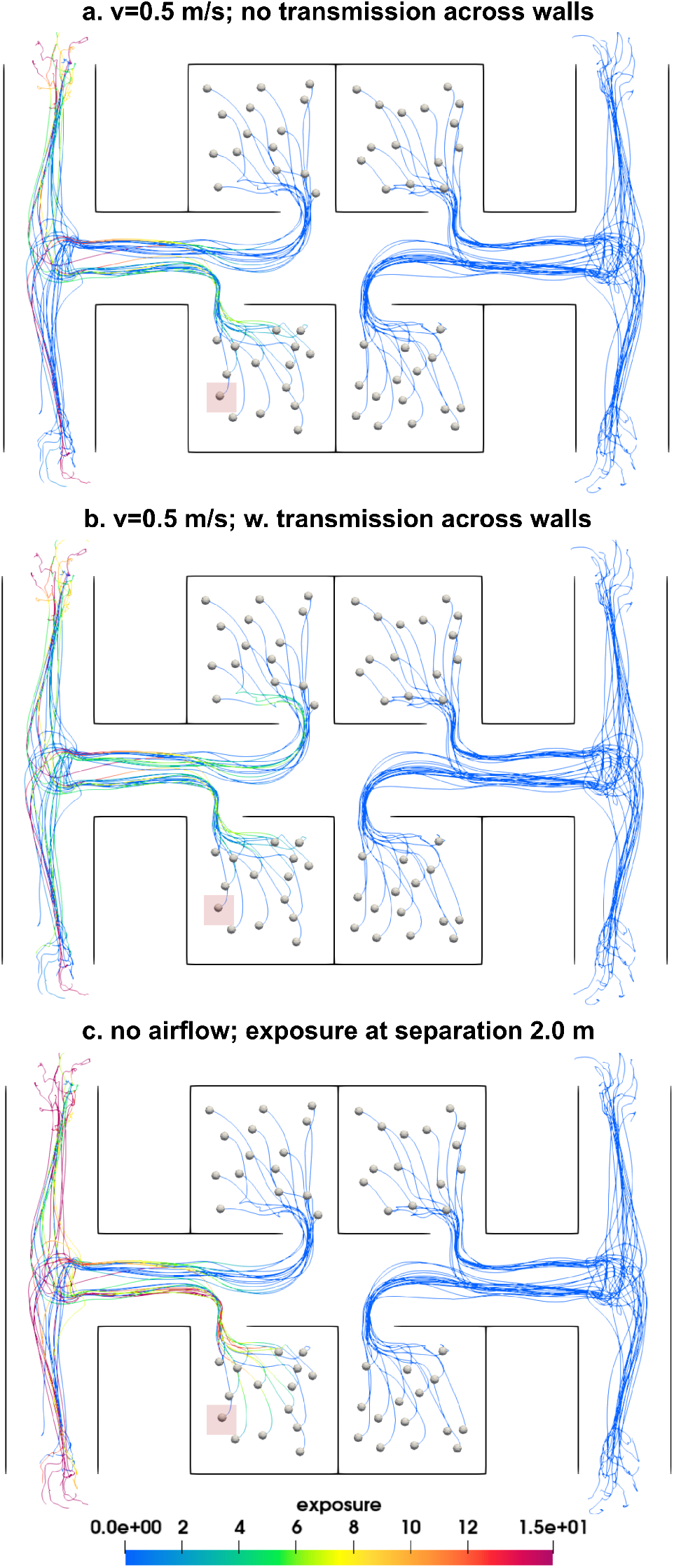
A comparison of exposure for flow velocity at 0.5 m/s without any transmission considered across boundary wall segments ℬ (panel a.); with transmission considered across boundary wall segments ℬ (panel b.); and for the scenario where exposure is estimated without considering airflow but with a fixed separation distance of 2.0 m (panel c.). Exposure is measured in terms of effective exposure duration, which is in the unit of time. The infected/index agent is marked in red square on all panels.

**Figure 7:**
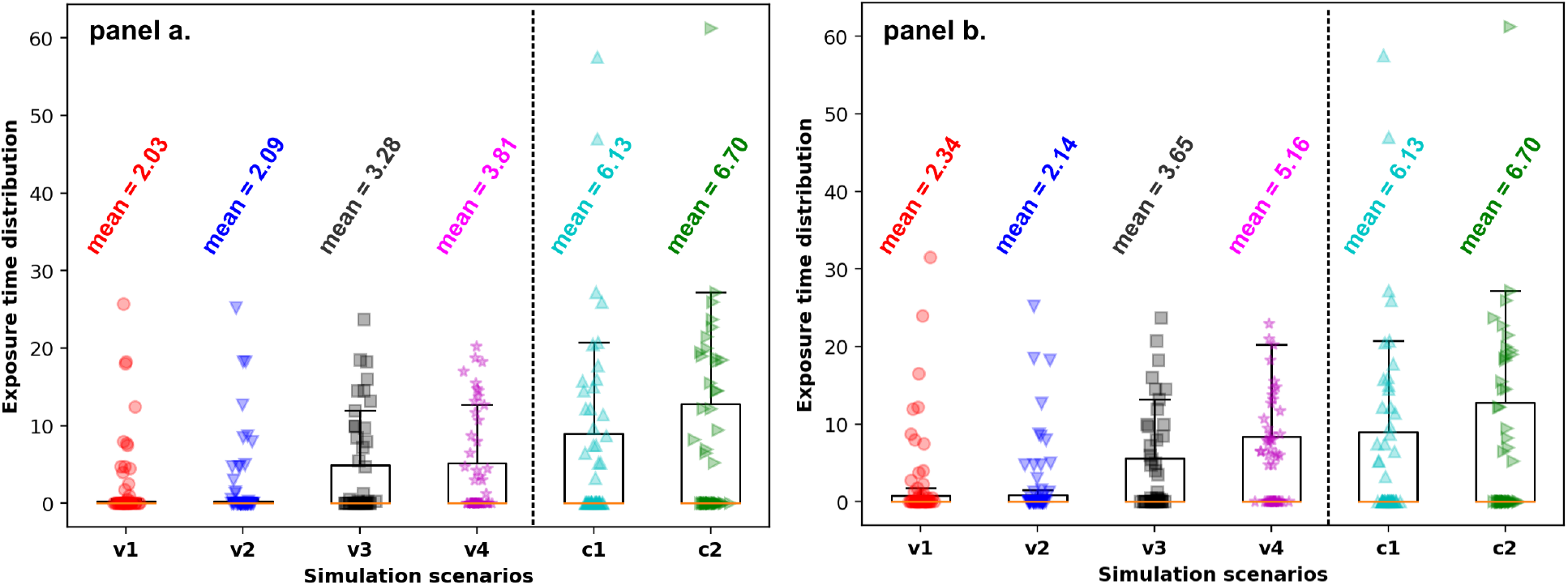
Comparison of agent exposure statistics for all N = 60 human occupant agents across all 12 simulation scenarios outlined in Section 5.2. The labels **v1, v2, v3, v4** denote the cases where airflow magnitude is varied as 0.1 m/s, 0.2 m/s, 0.5 m/s, and 1.0 m/s respectively. Labels **c1** and **c2** represent scenarios were separation distance threshold of 1.5 m and 2.0 m were employed to iedntify exposure. Cases without any transmission across wall boundary segments ℬ are on the left, while those with transmission across ℬ are on the right.

**Figure 8:**
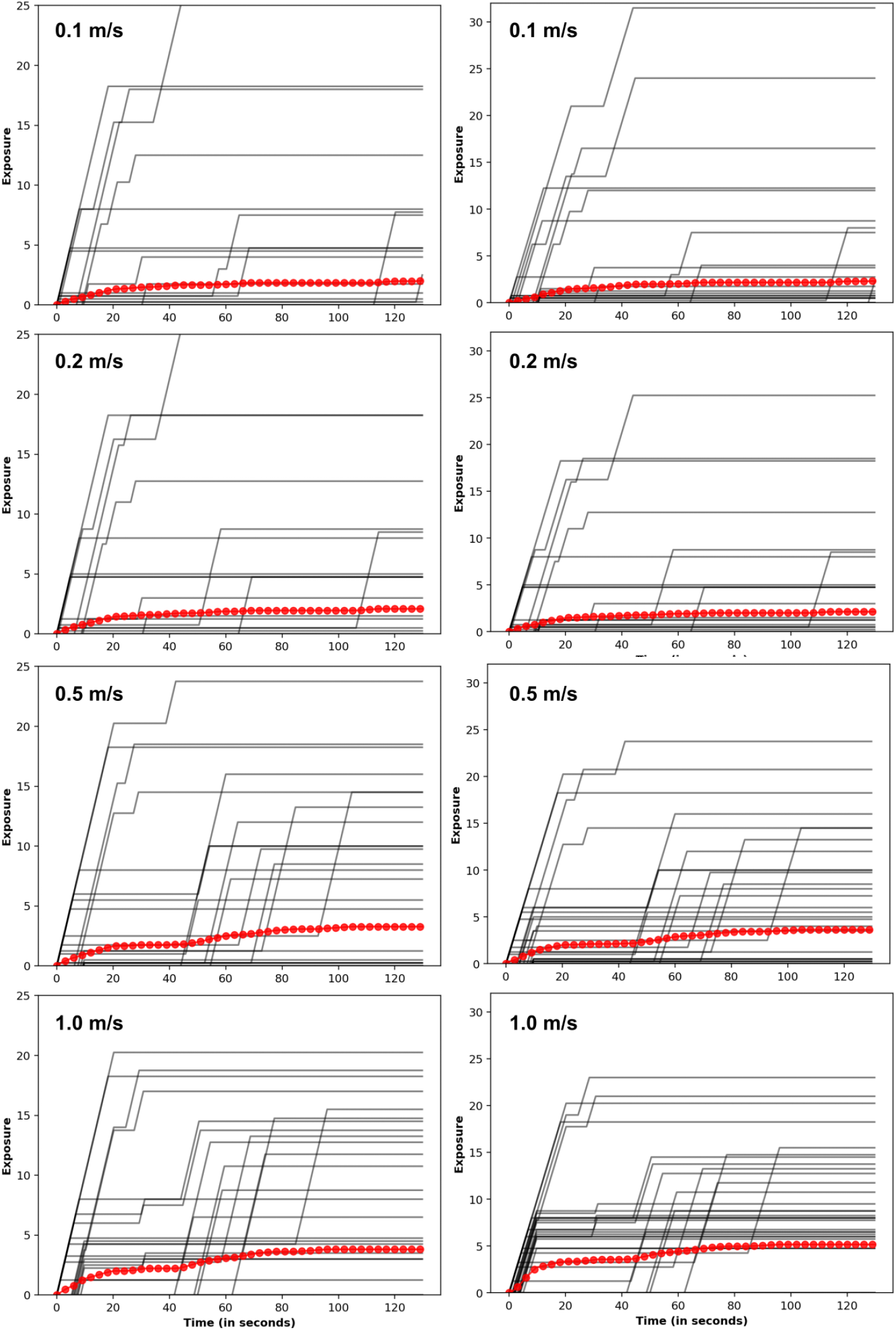
A compilation of exposure dynamics over time, for all 8 simulation scenarios with airflow based exposure models. Left column represents all cases without any transmission across boundary wall segments ℬ; while right column represents all cases with transmission across boundary wall segments ℬ. Airflow velocity magnitudes increase from top to bottom. Exposure is measured in terms of effective exposure duration, which is in the unit of time. Individual agent data is show in black, while agent ensemble averaged exposure dynamics curve is shown in red.

## 7 Discussion

Here, we have outlined the computational framework for modeling flow-mediated infection transmission in an indoor space amongst socially interacting human occupants. To the best of our knowledge, this is amongst the first instances of mesoscale transmission models that integrate flow physics and respiratory droplet transport considerations with active agent-based model for human occupant behavior in a computationally inexpensive manner. While we have demonstrated the model development and application for representative case-studies; several factors and assumptions need further discussion. First, we have not discussed here the computation of an actual infection probability or risk, but instead presented our analysis in terms of contact and exposure as surrogate parameters that correlate with infection. In practice, the contact and exposure data can be further used to estimate more detailed infection probabilities based on Wells-Riley equations and dose-response theory as proposed in^[52,62,7,18]^, and based on realistic pathogen-specific data such as viral shedding, viral load, and survivability^[67,56,9]^. Additionally, all agents are assumed to be equally susceptible here. In practice, agents may have varying degrees of susceptibility. For example, agents recently infected may have acquired immunity reducing susceptibility; similarly for vaccinated and masked individuals. This would require further advancement of the current model, where exposure leads to infection probabilities only for those agents marked susceptible; similar to the premise of SIR-type compartmental epidemiological models. Second, both droplet evaporation model and droplet dynamics model can be further advanced by incorporating higher-resolution respiratory droplet physics models^[4,10]^ to enable more accurate representation of respiratory droplet phenomena.

A discussion on what the airflow field represents in a top-down planar view of the room is also important in this context. Indoor airflow patterns are inherently 3-dimensional, and room height can factor in based on aspects such as location of inlet and exhaust vents, doors, and windows^[44,73]^. We note that averaging of the flow-field along agent height, for a representative mean height, will yield a physically representative depth-averaged flow-field that can be used as an input to the modeling framework with spatiotemporal interpolation techniques as identified in Section 4.4. Such approaches have been demonstrated in other well resolved coupled flow physics models^[36,39]^, where flow variables are interpolated at agent location on a planar grid. Such a description will capture the dominant large scale flow features in the room which will have the leading influence in determining the flow-mediated transmission dynamics. Avenues to integrate finer-scale flow features such as the human convective boundary layer^[34,35]^, and effect of local flow diverters (fans, blowers etc.) need to be further investigated for continuing advancement of the model framework. Additionally, the current model computes exposure based on droplet distance for all droplets, irrespective of where their vertical location (depth-wise in the room, *x*_2_ as per Section 3.2) is. This enables a sufficiently conservative exposure estimate, as droplets that may have fallen below the face location of susceptible individuals are included in exposure, allowing for some re-suspension of these droplets in the depth-wise direction due to strong flow patterns in/out of the simulation plane^[73]^. Future advancements of this mesoscale modeling approach will comprise additional investigations on these aspects, and systematic integration with complex building scale CFD data.

Another key aspect pertains to simulation parameters and their estimation. The framework encompasses several parameters and model choices, including: potential choices for ℛ, 𝒲, and 𝒜, and their associated parameters and constants; relaxation time; model constants for the social force model; droplet initial velocity and size; integration time *T*_*c*_ etc. Existing studies with social force simulations have demonstrated the feasibility of conducting large-scale parameter sweep studies^[49]^; and integrated simulation predictions with real-world human movement data and camera or closed-circuit images to do inverse parameter estimates^[60,3]^. Similar approaches can be employed using this modeling framework as well, given the relatively moderate computational cost for these models even as the number of agents increase. In the context of infection transmission, model predictions can also be integrated with contact tracing data^[28,12]^ to identify model parameter combinations. Lastly, parameter selection process can also be set-up using surrogate neural network based optimization models for a given building scenario, based on infection records for occupants of that building. In summary, despite the wide range of model parameters, several avenues exist for handling the parameter estimation problem; and will comprise another area of future model development and investigation.

## 8 Concluding remarks

While the Covid-19 pandemic has had an unprecedented impact on the world population, similar viral outbreaks have occurred before, and are likely to recur in the future to varying extents. The need for suitable quantitative models for understanding flow-mediated infection transmission; critically evaluating common assumptions such as 6-feet social distancing and well-mixed airflow; and planning control strategies and interventions remains timely and critical. We have devised and outlined a mesoscale model for flow-mediated infection transmission amongst socially interacting human occupants in an indoor space. This is amongst the first attempts to pair active agent-based models with flow physics models for respiratory infections. We have outlined various numerical and algorithmic details, and demonstrated model utility using a representative case-study. Data from the case-study clearly elucidates that accounting for airflow can lead to different infection transmission patterns compared to cases where no airflow information is considered; and that out model can capture this flow-mediated modality suitably. We emphasize the importance of analyzing flow-mediated infection transmission as an inherently multiscale problem; and in that context, our mesoscale model can complement finer scale respiratory droplet and airflow investigations, and larger scale epidemiological models. Continued investigations can enable using such models for devising scientific evidence-based guidelines for infection prevention and control during operations in facilities with human occupied indoor spaces.

## Supporting information

Supplemental animation: animation-01-lanes.mp4

Supplemental animation: animation-02-agents.mp4

Supplemental animation: animation-03-exposure.mp4

## Data Availability

All data produced in the present work are contained in the manuscript.

## 9 Conflicts of interest

Authors declare no conflicts of interest related to the content presented in this manuscript.

## 10 Acknowledgements

This work was conducted as part of a remote research internship arrangement for GW under supervision by DM. DM conceptualized the complete modeling framework with integration of droplet physics; devised the case-studies; developed post-processing code; analyzed the data; and drafted the manuscript. GW implemented the modeling framework into simulation codes and scripts; designed code integration with social force model; and ran all case studies involved. Both DM and GW reviewed and approved the final manuscript for publication. We thank Professor Shelly Miller from the University of Colorado Boulder for many valuable discussions regarding infection transmission. We also thank student researchers Akshita Sahni, Lindsey Nast, and Joseph Wilson, from the University of Colorado Boulder, for their assistance provided to GW during the course of this study.

## Figures and Tables

### Algorithm 1

An illustration of a complete algorithmic implementation of the overall computational framework, presented here for a given pre-computed airflow velocity dataset **<u>u</u>** with an explicit Runge-Kutta time integration scheme with Butcher array representation: 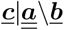.

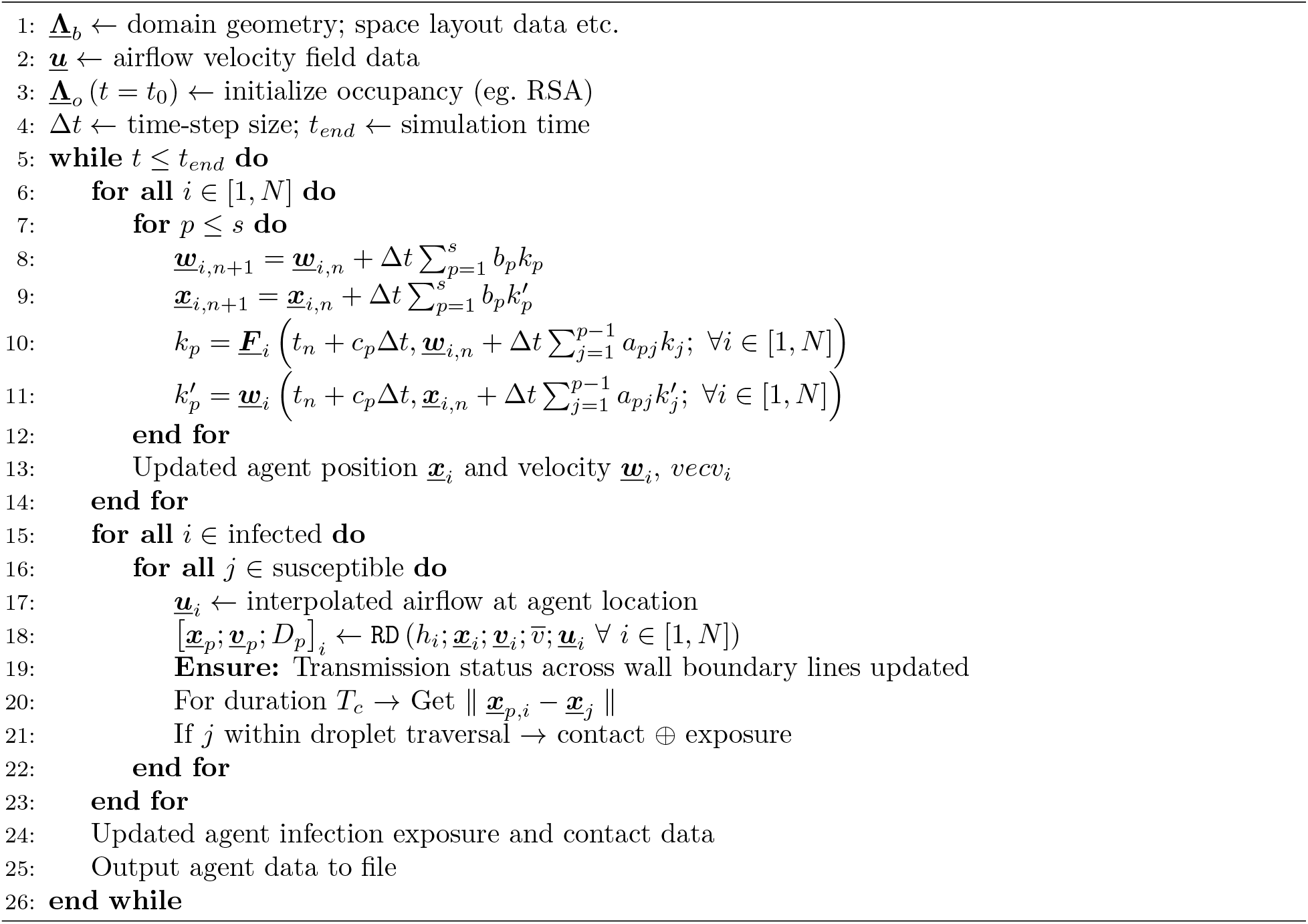

## Supplementary Material Information

Supplementary information for this manuscript is provided in form of additional visualization of simulation data as animations. Specifically, a set of three different animations have been provided here. First, animation movie file named: ***animation-01-lanes*.*mp4*** illustrates one of the sample simulations presented for validation of the social force model as outlined in Section 5.1, showing the spontaneous formation of lanes as demonstrated in Figure 2 in the main manuscript. Agents entering the walkway from left and right are distinguished in color red and yellow respectively. Second, animation movie file named: ***animation-02-agents*.*mp4*** illustrates the agent dynamics for the sample indoor space case study presented in Section 6, from each of their respective rooms to the exit doorways, as illustrated using snapshots in Figure 5. Third, animation movie file named: ***animation-03-exposure*.*mp4*** illustrates the dynamics of the exposure patterns mapped along the pathlines for each agent, for the three cases depicted in Figure 6. The second and third animations are sped up to a frame rate of 32 fps, to illustrate the dynamics from the room to the exit doorways in 16 seconds of view time.

## Notes

### Competing Interest Statement

The authors have declared no competing interest.

### Funding Statement

This study did not receive any funding

### Summary of Updates

This version has been updated to include updated discussion of literature, specifically placing the methodology in our work in the context of other methodologies in terms of computational expense. Several mathematical typographical errors have also been corrected. Lastly, a couple of figure captions have been updated to clarify that exposure in our paper is measured in units of time.

